# Potent anti-SARS-CoV-2 Antibody Responses are Associated with Better Prognosis in Hospital Inpatient COVID-19 Disease

**DOI:** 10.1101/2020.08.22.20176834

**Authors:** Patrick J. Tighe, Richard A. Urbanowicz, C. Lucy Fairclough, C. Patrick McClure, Brian J. Thomson, Nancy Gomez, Joseph G. Chappell, Theocharis Tsoleridis, Matthew Loose, Matthew Carlile, Christopher Moore, Nadine Holmes, Fei Sang, Divyateja Hrushikesh, Gemma Clark, Nigel Temperton, Tim Brooks, Jonathan K. Ball, William L. Irving, Alexander W. Tarr

## Abstract

COVID-19 continues to cause a pandemic, having infected more than 20 million people globally. Successful elimination of the SARS-CoV-2 virus will require an effective vaccine. However, the immune correlates of infection are currently poorly understood. While neutralizing antibodies are believed to be essential for protection against infection, the contribution of the neutralizing antibody response to resolution of SARS-CoV-2 infection has not yet been defined. In this study the antibody responses to the SARS-CoV-2 spike protein and nucleocapsid proteins were investigated in a UK patient cohort, using optimised immunoassays and a retrovirus-based pseudotype entry assay. It was discovered that in severe COVID-19 infections an early antibody response to both antigens was associated with improved prognosis of infection. While not all SARS-CoV-2-reactive sera were found to possess neutralizing antibodies, neutralizing potency of sera was found to be greater in patients who went on to resolve infection, compared with those that died from COVID-19. Furthermore, viral genetic variation in spike protein was found to influence the production of neutralizing antibodies. Infection with the recently described spike protein variant 614G produced higher levels of neutralizing antibodies when compared to viruses possessing the 614D variant. These findings support the assertion that vaccines targeting generation of neutralizing antibodies may be useful at limiting SARS-CoV-2 infection. Assessment of the antibody responses to SARS-CoV-2 at time of diagnosis will be a useful addition to the diagnostic toolkit, enabling stratification of clinical intervention for severe COVID-19 disease.

## Introduction

The current COVID-19 pandemic caused by SARS-CoV-2 infection [1] has highlighted the potential for high rates of transmission and mortality from this coronavirus in an immune naïve global population. As of 13 August 2020 there have been ∼20 million confirmed infections worldwide, with more than 750,000 deaths associated with severe COVID-19 disease [2]. The long-term impact of this infection and the potential for post-infection sequelae are still poorly understood. Understanding the protective and pathogenic consequences of the immune response to this infection, the potential markers for different disease outcomes, and the potential correlates of protection are essential for clinical management of SARS-CoV-2 infection and future vaccine development.

Coronavirus infections are associated with potent antibody responses that are believed to persist after infection (reviewed in [3]). MERS CoV infections resulting in pneumonia were strongly associated with prolonged seropositivity [4], with antibodies appearing three weeks after onset of illness [5]. However, in MERS infections that result in mild symptoms, antibodies were only present transiently [6]. Surveillance of SARS-CoV-2 infections has relied on a combination of RTPCR tests and serologic assays. ELISA assays detecting antibodies binding to the nucleocapsid protein (N) and spike antigen (S) have found widespread use, with both the S1 subunit of spike and the receptor binding domain (RBD) being used as targets for antibody detection [7]. Lateral flow assay (LFA) devices have also been implemented for detection of antibodies, but the sensitivity of these serology tests in early infection has been questioned [8]. In addition, it appears that some SARS-CoV-2 infections result in no antibodies generated to the reference antigens used in these assays [9]. In cases where anti-SARS-CoV-2 antibodies are generated, circulating antibodies wane within months [10]. Antibody production may be a useful marker of a successful immune response to SARS-CoV-2, but the role of antibodies in disease pathogenesis is controversial. It has been reported that generation of IgG specific for the RBD is associated with poor clinical outcomes [11].

Sequencing of the SARS-CoV-2 genomes has revealed eleven SARS-CoV-2 genetic lineages, whose prevalence differ between geographical location [12]. It is still unclear what influence genetic polymorphisms have on the antigenicity of infections. While genetic variation in the spike protein is limited, a polymorphic site has been identified at amino acid 614 [13]. This amino acid is associated with differing ability of the virus to shed the S1 subunit. While aspartic acid is the most common amino acid at this position, glycine is also observed, and believed to result in stabilised retention of spike on the virus surface [14] and improved infectivity in cell culture [13].

In this study we assessed antibody reactivity to the S1 subunit of spike protein and the nucleocapsid protein of SARS-CoV-2 in hospitalised patients with different clinical outcomes, with the aim of determining if antibody responses, assessed as either sero-reactivity or neutralization potency, were associated with COVID-19 disease progression. We also interrogated the influence of the Spike D614G polymorphism on antibody responses during infection in COVID-19-confirmed cases to determine if aa614 influenced immunogenicity.

## Methods

### Clinical samples

422 serum samples were obtained from 227 different hospital in-patients with symptomatic RT-PCR proven COVID-19 infection. These samples were diagnostic specimens collected for routine clinical chemistry analyses. Surplus material was provided anonymously for development of novel serology and virus neutralization assays. Review by the School of Life Sciences Ethical Review Committee deemed the study to not require full ethical review. Patients were predominantly of white British ethnicity and the majority were elderly (Table 1).

**Table 1:** Characteristics of the study population analysed in this study

|  | **DISCHARGED** |  | **DIED** |  |
| --- | --- | --- | --- | --- |
|  | N | % | n | % |
| **MEDIAN AGE** | 74 |  | 82 |  |
| **MALE** | 73 | 52.52 | 45 | 52.33 |
| **FEMALE** | 66 | 47.48 | 41 | 47.67 |
| **ETHNICITY** |  |  |  |  |
| **WHITE** | 88 | 63.31 | 68 | 79.07 |
| **BLACK** | 7 | 5.04 | 1 | 1.16 |
| **ASIAN** | 9 | 6.47 | 0 | 0.00 |
| **MIXED** | 1 | 0.72 | 1 | 1.16 |
| **NA** | 34 | 24.46 | 16 | 18.60 |
| **ICU** | 42 | 30.22 | 18 | 20.93 |
| **NO ICU** | 96 | 69.06 | 68 | 79.07 |
| **ICU DATA N/A** | 1 | 0.72 | 0 | 0.00 |
| **MEDIAN DURATION OF STAY (D)** | 15 |  |  |  |
| **MEDIAN TIME TO DEATH (D)** |  |  | 11 |  |

Sequence data was generated as part of the data collection for the COVID-19 Genomics UK (COG-UK) Consortium, utilising extracted viral RNA from throat swabs generated for routine qRT-PCR analysis of inpatients (with approval from PHE Research Ethics and Governance Group (REGG) R&D NR0195). Anonymised clinical data for these patients was provided by clinical staff at Nottingham University Hospitals Trust. 192 sera collected in August 2019 as part of the Trent HCV cohort study [15], were assumed to be negative for anti-SARS-CoV-2 antibodies. Sera were inactivated before use using the WHO-approved protocol of treatment with 1% Triton X-100 for 4 hours before use in serological assays.

### Anti-S1 and anti-nucleocapsid ELISA

Serum samples were serially diluted in 3% skimmed milk powder in PBS containing 0.05% Tween 20 and 0.05% sodium azide. All assays were performed on Biotek Precision liquid handling robots in a class II microbiological safety cabinet. For endpoint dilution ELISAs, sera were progressively 4-fold diluted from 1:150 to 1;38,400. ELISA was performed by coating 384 well Maxisorp (NUNC) assay plates with either 20 µL per well of 0.5 µg.mL^−1^ of Wuhan strain SARS-CoV-2 spike protein S1 subunit (His tagged, HEK293 expressed; Sino Biological) or SARS-CoV-2 nucleocapsid (His Tagged, baculovirus expressed; Sino Biological) in carbonate-bicarbonate buffer (CBC; Merck), or human IgG at 1 µg.mL^−1^ in CBC buffer as controls. Plates were sealed with foil film and incubated overnight at 4 °C. Plates were then washed with PBS with 0.05% Tween 20 (PBS-T) 3 times using a ThermoFisher Wellwash Versa plate washing robot. Wells were immediately filled with 100 µL of 3% skimmed milk powder (w/v) in PBS and 0.05% sodium azide (PBS-MA) and blocked overnight at 4 °C. Assay plates were then washed 3 times and 20 µL of pre-diluted serum sample (including SARS-CoV-2 antibody-positive and negative serum controls) added in duplicate wells. After one hour at 21 °C, the plate was washed 3 times in PBS-T, followed by addition of 20 µL of gamma chain-specific anti-human IgG-HRP conjugate (Sigma A0170–1ML) at 1:30,000 dilution, incubating for one hour at 21 °C. Following a final three washes with PBS-T, 40 µL One-step UltraTMB substrate solution (ThermoScientific) was added to each well. After incubating for 20 minutes at room temperature, 40 µL of 2N H_2_SO_4_ was added to each well and Absorbance was measured at 450nm using a GlowMax Explorer microplate reader (Promega).

### Commercial anti-S1 and anti-N assays

Detection of antibodies to the S1 protein (Euroimmun) and N Protein (Roche) were performed at the Rare & Imported Pathogens Laboratory at Public Health England using the standard protocol.

### SARS-CoV-2 pseudotype neutralization assay

Pseudotypes were produced as essentially described previously for other viruses [16, 17]. Briefly, 1.5 × 10^6^ HEK293T cells were seeded overnight in a 10 cm diameter Primaria-coated dish (Corning). Transfections were performed with 2 µg each of the murine leukemia virus (MLV) Gag-Pol packaging vector (phCMV-5349), luciferase encoding reporter plasmid (pTG126) and plasmid encoding Wuhan strain SARS-CoV-2 spike using 24 µL polyethylenimine (Polysciences) in Optimem (Gibco). Reaction mixtures were replaced with Complete DMEM after 6 h. A no-envelope control (empty pseudotype) was used as a negative control in all experiments. Supernatants containing SARS-CoV-2 pseudotype viruses (pv) were harvested at 72 h post-transfection and filtered through 0.45 μm membranes. For infectivity and neutralization testing of SARS-CoV-2 pv, 2 × 10^4^ VeroE6 cells/well were plated in white 96-well tissue culture plates (Corning) and incubated overnight at 37 °C. The following day, SARS-CoV-2 pv-containing supernatants were incubated with sera (pre-treated with 1% Triton X-100/heat-inactivated for 1 hr at RT), before being added to VeroE6 cells for 4 h. Infection inocula were then discarded and 200 µL DMEM added to the cells. After 72 h, media was discarded, cells lysed with cell lysis buffer (Promega) and relative luciferase activity (in RLU) measured using a FLUOstar Omega plate reader (BMG Labtech). Each sample was tested in triplicate.

### Assessment of polymorphisms in the S protein

Sequence polymorphisms at the D614G site in spike were determined as part of the GISAID SARS-CoV-2 project data collection. Briefly, viral cDNA was synthesised using the RNA to cDNA EcoDry Premix (Random Hexamers) (Takarabio). Sequencing libraries were prepared from cDNA using the ARTIC amplicon sequencing protocol v2 [18] using the Oxford Nanopore Native Barcoding Ligation Sequencing protocol and the Native Barcoding Expansion 1–12 and 13–24 kits (Oxford Nanopore Technologies). After quantification using a Qubit 1X dsDNA HS Assay Kit (Thermo Fisher Scientific), 20 ng of library was loaded onto a MinION flow cell (Oxford Nanopore Technologies; FLO-MIN106 R9.4.1) and sequenced on the GridION X5 Mk1. Data was collected and analysed using RAMPART [19], as previously described [20]. Samples from our cohort included in the UK-wide sequence analysis were compared for reactivity in both serology and neutralization assays.

### Statistical analyses

All analyses were performed using GraphPad Prism version 8.4.3. Assessment of normal distributions was performed using D’Agostino & Pearson tests. Data generated by ELISA and neutralization assays was analysed using Mann-Whitney U Test for comparison of two sets of data. Spearman correlation coefficients were calculated for comparison of continuous datasets.

## Results

### Cross-sectional analysis of antibody responses to SARS-CoV-2 in hospitalised patients

ELISA assays were developed for quantitative assessment of antibodies directed to the SARS-CoV-2 Spike S1 subunit and the nucleocapsid proteins (Supplementary Figure 1). These data were qualitatively concordant with serology testing performed using CE-marked commercial ELISA assays on a subset of 24 samples taken from RT-PCR-confirmed COVID-19 cases (data not shown). Sampling of a cross-section of patients at different times following initial diagnosis of infection was performed to interrogate the timing of the antibody responses to S1 and N proteins during severe COVID-19. We tested 192 samples retrieved from 142 discrete patients based on time since diagnosis by RT-PCR. Sampling was performed from 4 days before diagnosis to 31 days after diagnosis. The majority of samples tested more than 14 days after diagnosis were found to possess antibodies to both the S1 (Figure 1A) and N proteins (Figure 1B). However, seven samples taken later than 14 days after diagnosis were negative for antibodies to both proteins. For some patients, sera were available shortly before the confirmatory RT-PCR test was performed. Antibody binding to N and S1 was observed for many of these samples, indicating that in some cases of severe COVID-19 disease the antibody response was already established at time of diagnosis. When directly comparing the anti-S1 and anti-N response in these samples it was observed that in the majority of cases anti-N antibodies were present if antibodies to the S1 spike subunit had been generated (Figure 1C).

**Figure 1.**
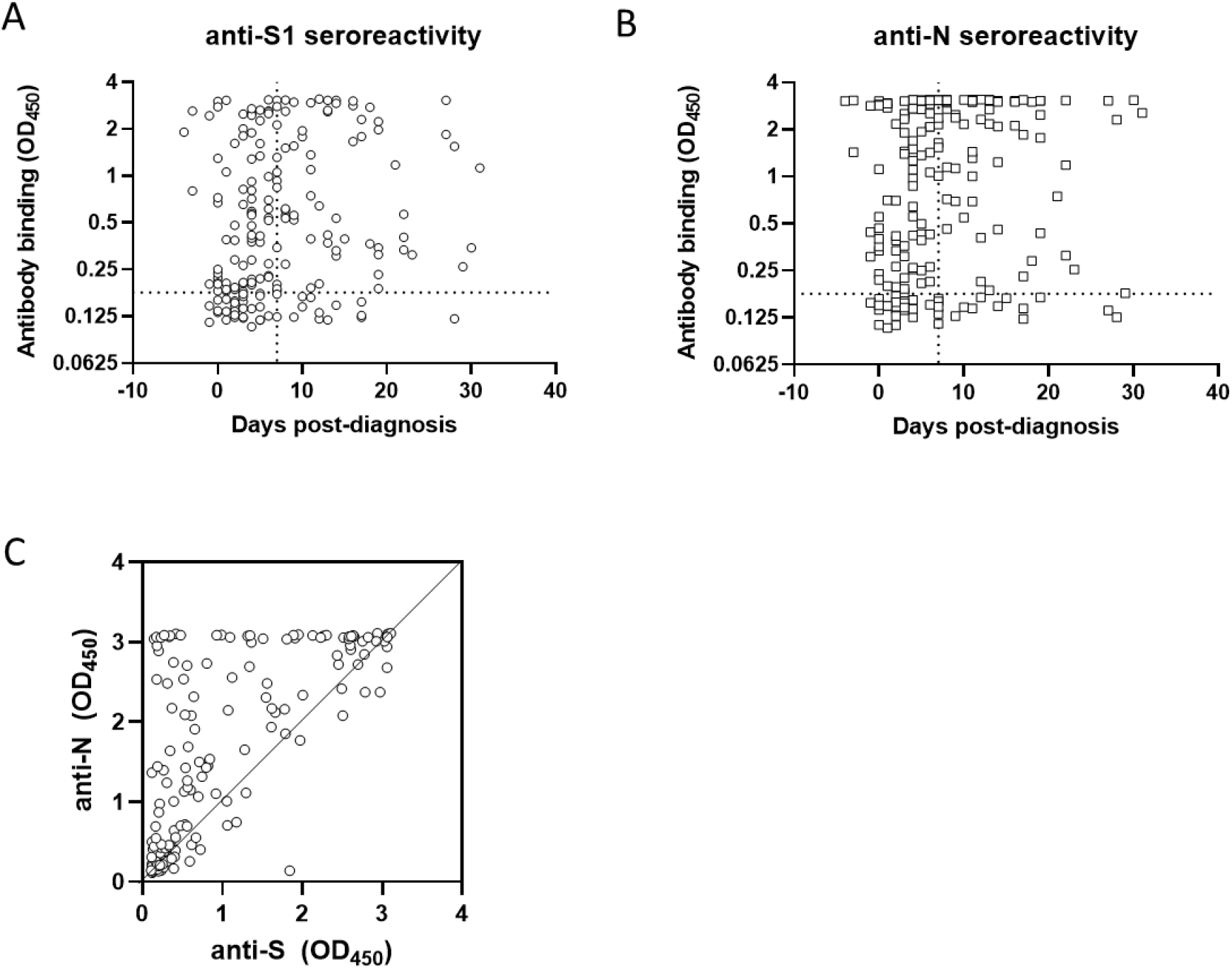
Development of the antibody responses to SARS-CoV-2 spike S1 and nucleocapsid proteins in hospital inpatients. ELISA using 192 samples taken at different times from initial diagnosis of infection by RT-PCR, performed using two viral antigens. Antibodies to A) spike S1 and B) N were detected for most sample following 7 days of diagnosed infection (indicated by a vertical dashed line). Assay threshold is indicated by a horizonal dashed line. C) Comparison of anti-S1 and anti-N signal for each sample. For clarity, equal reactivity is indicated by the diagonal line.

### Antibody response is associated with outcome of infection

To investigate whether the antibody response to SARS-CoV-2 was associated with recovery in this patient group, sero-reactivity data (defined as the greatest binding signal observed for each patient) and age were compared according to outcome. As expected, patients who died during hospitalisation with COVID-19 were more likely to be older (median age at diagnosis was 74.2 y for discharged patients versus 82.3 y for patients who died). Antibody responses detected in samples collected after 7 days following diagnosis were associated with age. Very young patients, and the very elderly, mounted a reduced antibody response to both S1 (Figure 2B) and nucleocapsid (Figure 2C). When samples taken between 7 days and 31 days following diagnosis were selected only from patients > 70 years old, a significant difference was observed in signal achieved in patients who survived infection and were discharged, and those who died during their inpatient stay (Figure 2D).

**Figure 2.**
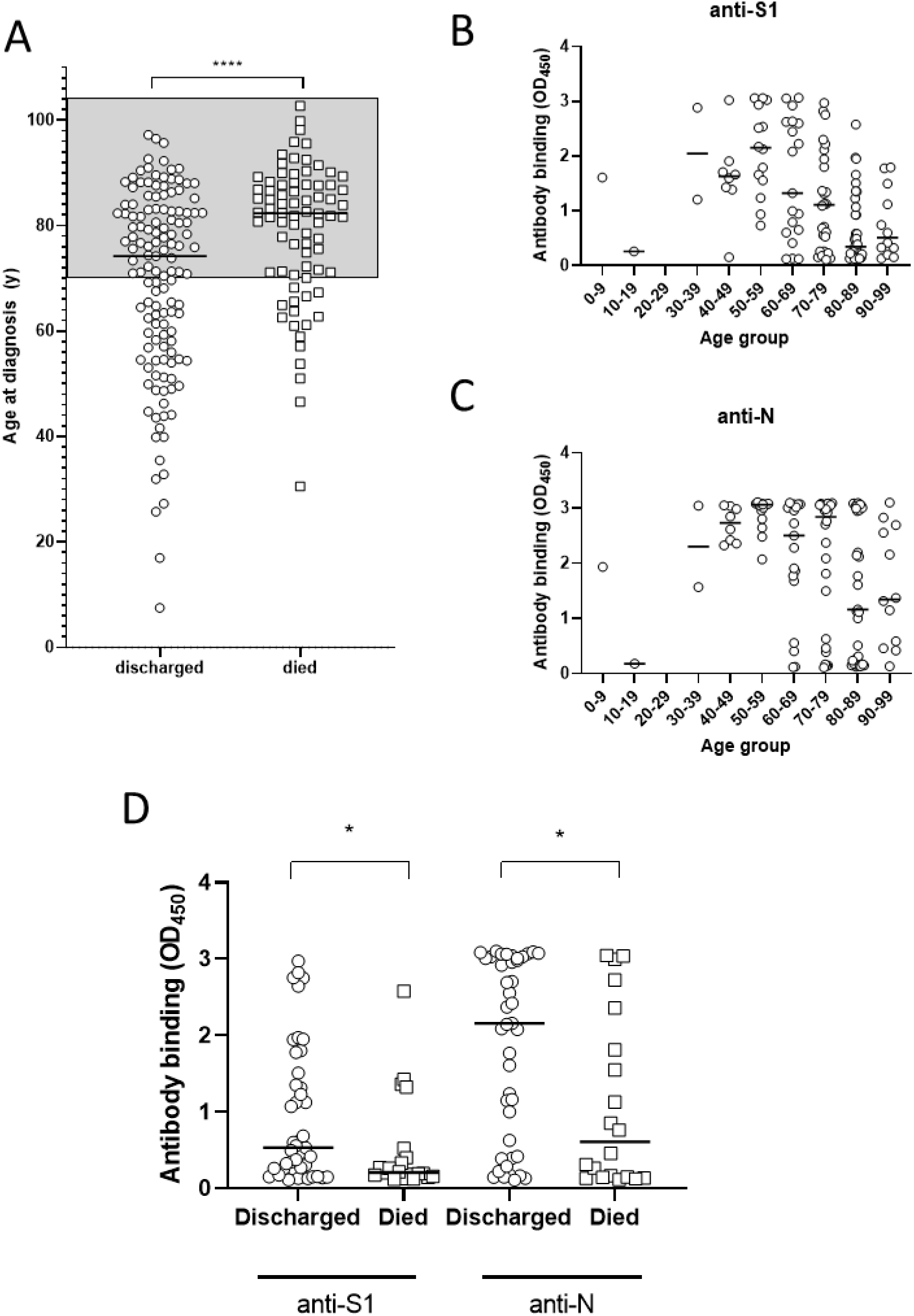
Association of the anti-spike S1 and anti-N antibody responses with age and outcome of infection. A) The median age of patients who survived infection and were discharged from hospital (n = 131) was lower than those patients who died of Covid-19 (n = 78) The shaded area highlights the patients that were selected for analysis in Figure 2 D. Maximal antibody responses in 120 discrete patients to B) S1 and C) nucleocapsid were associated with age, with younger and older patients producing the weakest antibody responses to both antigens. D) Selecting patients > 70 years old, antibody responses in samples between 7 and 31 days after diagnosis were assessed to both antigens. **** p< 0.001 *p>0.05; Mann-Whitney U test.

### An early potent antibody response to S1 and N is associated with outcome of infection

Having identified differences in the antibody responses associated with outcome, the difference in antibody response early in severe infection was analysed. A total of 149 discrete patients with a sample available within the first seven days of diagnosed infection were analysed (86 who were subsequently discharged and 63 who died as an inpatient) (Figure 3). There was no significant difference in sampling times for these groups (Figure 3A). Comparing the maximal binding signal achieved for these patients in the first 7 days following diagnosis, greater median reactivity in both anti-S1 and anti-N assays was achieved for patients who subsequently cleared infection (Figure 3B). As saturation in our assays was observed for a small number of the samples analysed, endpoint titrations were performed for each sample. Some samples did not achieve signal above background and were omitted from this analysis. Endpoint titration and single point signals were highly correlated for both the anti-S1 assay (Spearman r = 0.6908, p< 0.0001) (Figure 3C) and the anti-N assay (Spearman r = 0.8771, p< 0.0001 (Figure 3D). Comparing endpoint titration for each sample in this analysis, both anti-S1 (Figure 3E) and anti-N antibody (Figure 3F) titres were greater in patients who were subsequently discharged following resolution of infection.

**Figure 3.**
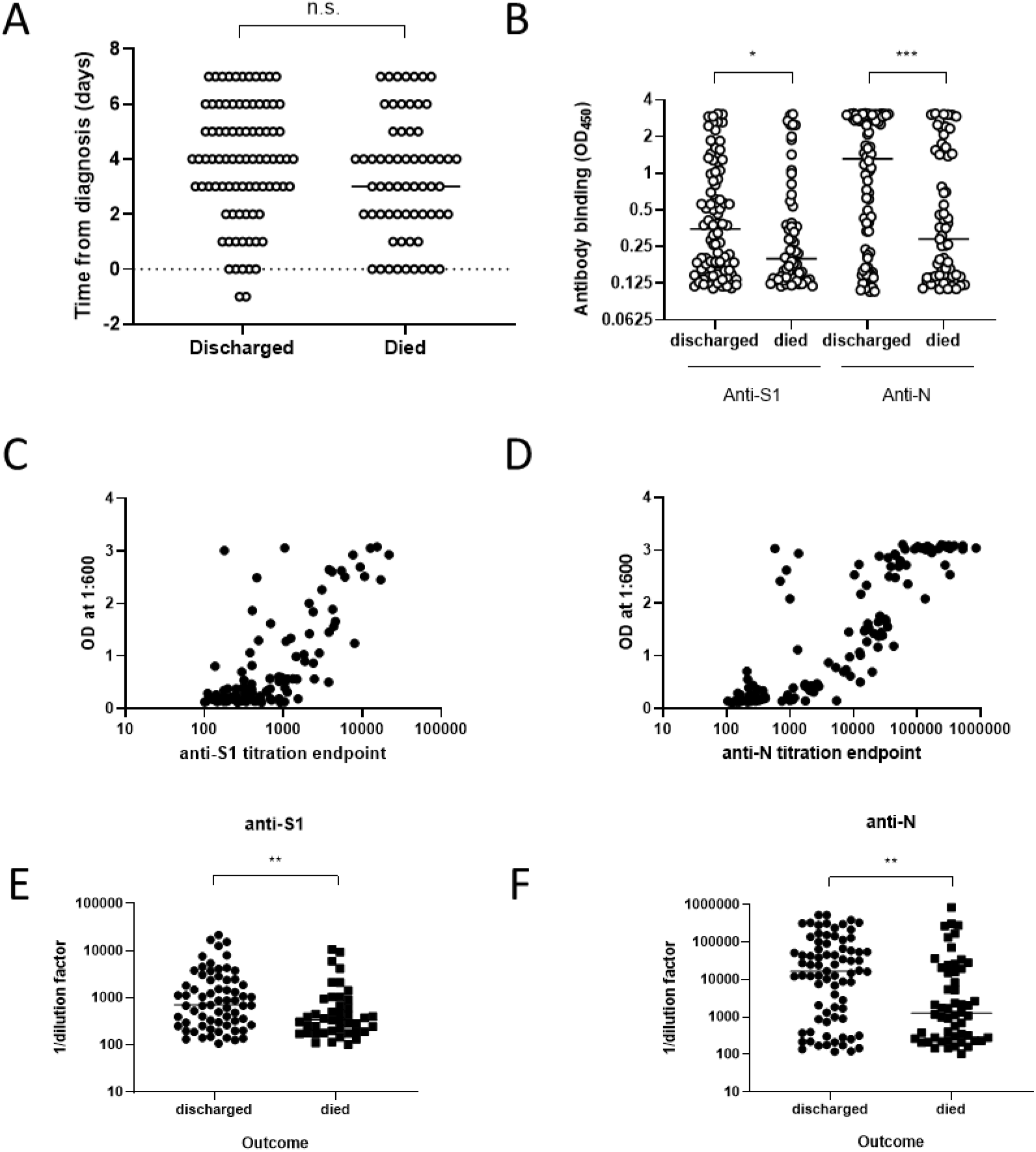
Early antibody response in hospitalised individuals is associated with improved Covid-19 prognosis. A) Sampling times up to day 7 following diagnosis for 86 recovering patients and 63 patients who died as a hospital inpatient following SARS-CoV-2 infection. B) Comparison of the ELISA signal in samples taken up to day 7 following diagnosis to both S1 and N proteins in patients with differing outcome. C) Comparison of the fixed-dose signal and endpoint titration for anti-S antibodies (Spearman r = 0.6908, p< 0.0001). D) Comparison of the fixed-dose signal and endpoint titration for anti-N antibodies (Spearman r = 0.8771, p< 0.0001). E) Anti-S1 endpoint titres in samples taken up to day 7 in patients who resolved infection or died from Covid-19. F) Anti-N endpoint titres in the first week of diagnosis in patients who resolved infection or died from Covid-19. Comparisons were performed using Mann Whitney U test. *p< 0.05 **p< 0.01

### Neutralizing capacity of serum antibodies is not strongly predicted by anti-S1 or anti-N sero-reactivity

Sera from our cohort were tested in a SARS-CoV-2 neutralization assay, utilising retroviral pseudotypes possessing the spike proteins of the Wuhan strain of SARS-CoV-2. A broad range of neutralization potency was observed, with reproducible neutralization data achieved in this assay (Figure 4A). The neutralization potency of sera taken at different times after diagnosis was analysed. While the majority of patients produced neutralizing antibodies within the first 10 days following diagnosis, some sera reproducibly enhanced infection, with up to 60% enhancement in signal observed (Figure 4B). When plotted against ELISA binding signal, a significant positive correlation was observed between neutralization with antibody binding to either S1 (Spearman r = 0.447; p< 0.0001; Figure 4C) or N (Spearman r = 0.5285; p< 0.0001; Figure 4D). A potent neutralizing antibody response produced during the first 7 days following diagnosis by RT-PCR was associated with better prognosis, with patients who survived COVID-19 disease having more potently neutralizing serum antibody responses than those who died (Figure 4E).

**Figure 4.**
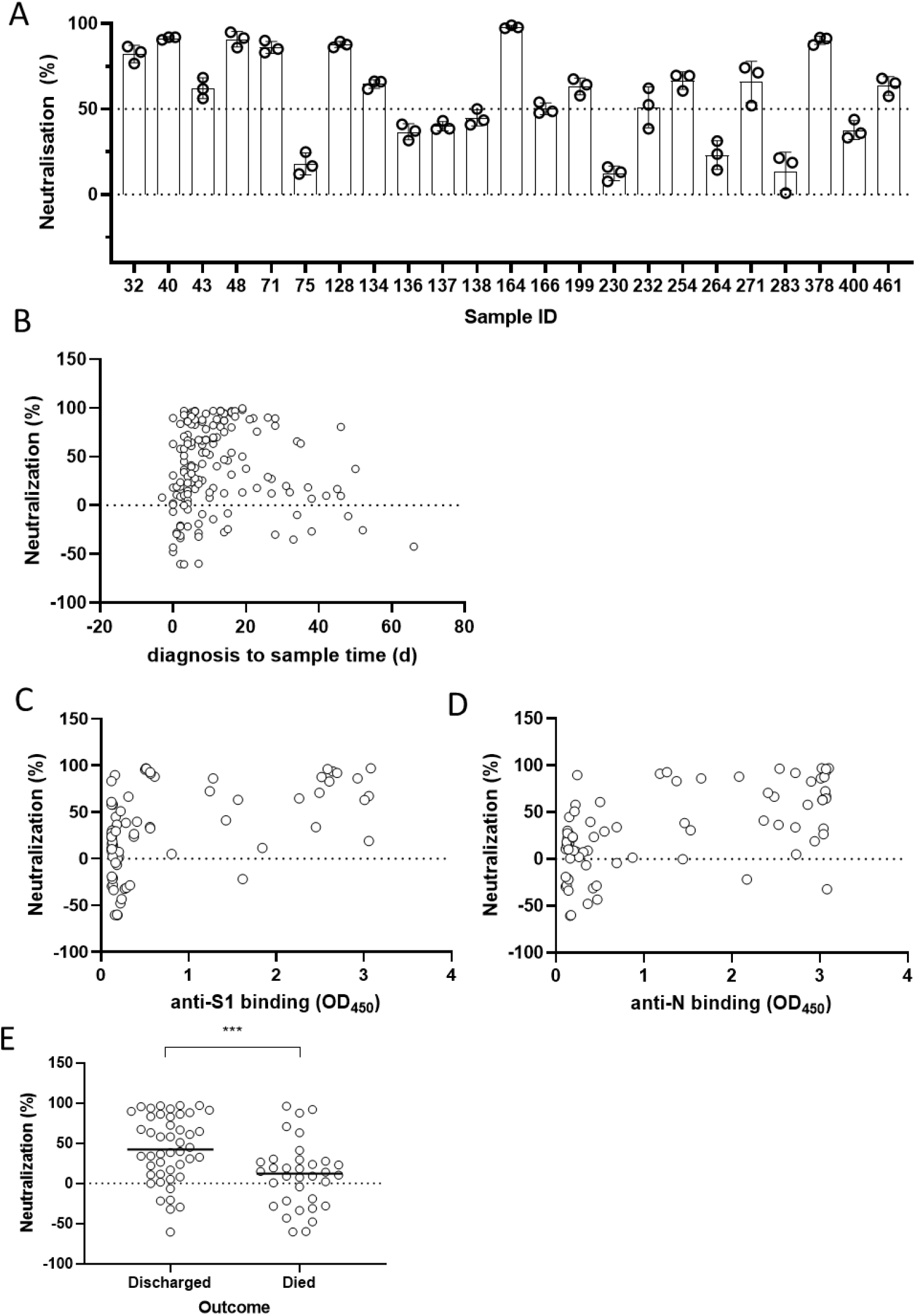
Neutralizing antibody responses in severe COVID-19 infections. A) Differing neutralization potency of sera from SARS-CoV-2 infections for SARS-CoV-2pp. All assays were performed in triplicate. This dataset highlights the intra-assay reproducibility of observed neutralisation. B) Neutralization potency of sera plotted against time from diagnosis by RT-PCR (n = 167). C) & D) Neutralization potency of 80 sera from independent patients sampled up to day 7 following diagnosis was compared to C) anti-S1 reactivity (Spearman r = 0.447; p< 0.0001) and D) anti-nucleocapsid reactivity (Spearman r = 0.5285; p< 0.0001). E) Neutralizing potency of sera taken up to day 7 following diagnosis, grouped by infection outcome. *** p< 0.001, Mann Whitney Test

### Polymorphisms in the spike gene are associated with generation of the neutralizing antibody response in severe COVID-19 disease

It has recently been described that a single nucleotide substitution leading to a replacement of aspartate with glycine residue at amino acid position 614 in the spike protein has arisen during transmission of SARS-CoV-2 infection, to become highly prevalent in some geographic locations, including Europe [13]. To investigate if this polymorphism was associated with differences in the generation of antibodies during infection in hospitalised patients, whole-genome SARS-CoV-2 sequencing data was generated for a subpopulation of our cohort where suitable swab samples had been taken for analysis for the UK COG consortium sequencing project [21]. Sequencing data was available for 100 patients where we had already generated antibody serology data. Choosing the timepoint for each patient that had the greatest signal in the anti-S1 ELISA, binding data was stratified by the D614G polymorphism (Figure 5). While there was no significant difference in the reactivity to S1 in ELISA in these two groups (Figure 5A), antibody responses to nucleocapsid protein were significantly higher in the G614-infected population (Figure 5B). Importantly, the neutralization potency observed in patients infected with the G614 variant was significantly higher than those infected with the D614 variant (Figure 6C).

**Figure 5.**
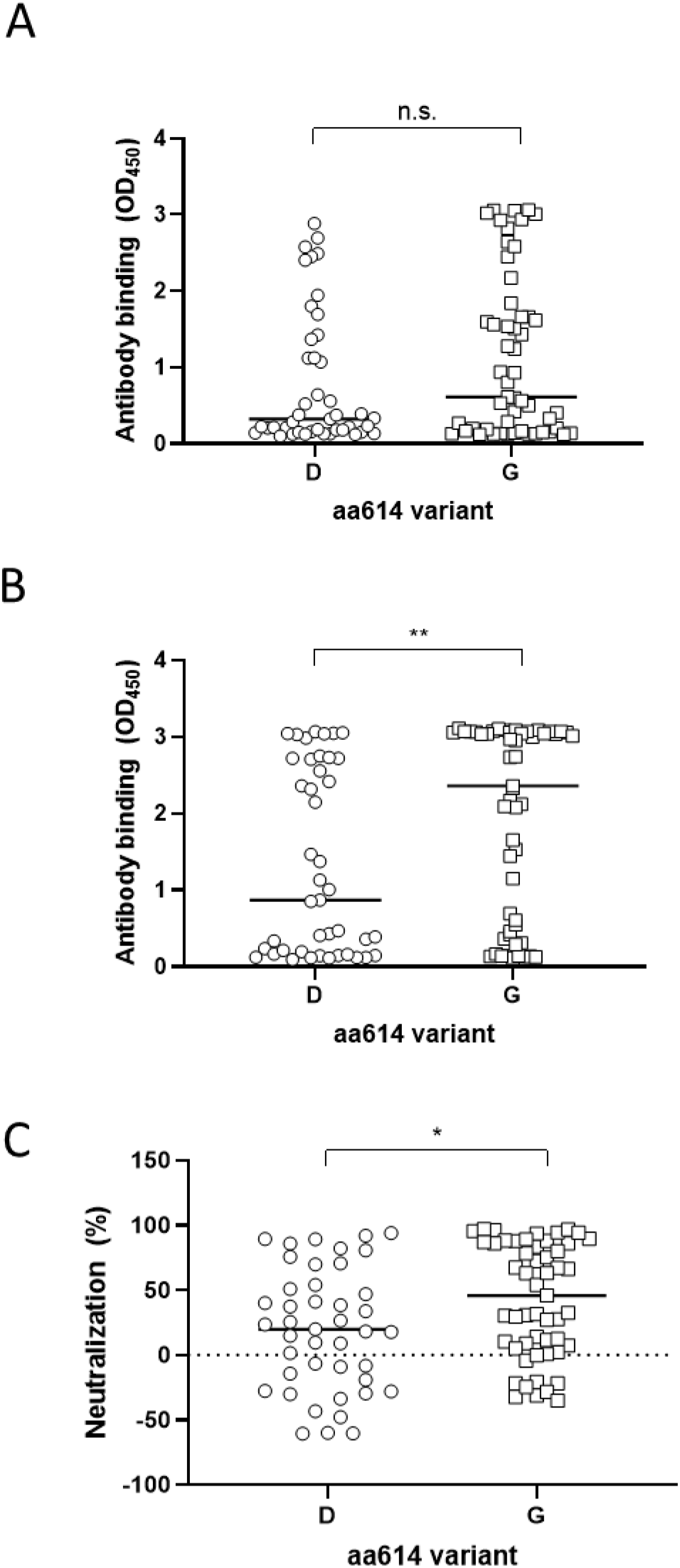
Association of polymorphisms in the spike protein on antibody reactivity and neutralization. Peak ELISA reactivity to A) S1 and B) N in 100 discrete patients with available viral genome sequence data was grouped based on the D614G polymorphism in the S1 subunit of spike protein, irrespective of disease outcome. C) Neutralization potency of sera in 96 discrete patients infected with viruses possessing the Spike 614D or 614G variants. Median values are plotted in each case. Medians were compared using Mann Whitney Test. * p< 0.05; ** p< 0.01; n.s. not significant.

## Discussion

For the first time, we have shown that, in hospitalised patients, the induction of an early high titre antibody response is associated with recovery from COVID-19 infection. While this contrasts with previous analysis of antibody response in severe COVID-19 infections [11], our optimisation of the quantitative resolution of antibody assays may provide better insight than data achieved in existing clinical assays that utilize saturating amounts of serum antibodies for detection. In our assay a correlation was observed between the interpretation of our fixed dilution ELISA and assays reporting the serially-diluted endpoint, and as such provide an accurate assessment of the antibody binding to the S1 subunit of spike and the nucleocapsid protein. These new assays may contribute to patient management and may assist with understanding the vaccine-induced antibody responses that are important for protection against severe COVID-19.

The association between the presence of antiviral antibodies and better prognosis in our mainly white, British cohort is striking, but needs to be confirmed in other cohorts. Consistent with other recent studies, antibodies were demonstrated to occur in the majority of patients a week after diagnosis [22, 23]. However, investigation of the early antibody response where present revealed that those individuals who mounted an IgG antibody response in the first week following diagnosis were more likely to have a positive prognosis. A study investigating antibodies directed only to the spike RBD identified an association between antibody production and poor prognosis in a Chinese cohort [11]. It may be that antibody specificity is important for determining protection, as demonstrated for other viruses [24]. While we did not investigate the IgM response in these infections, analysis of the antibody repertoire and IgM/IgG subclasses may provide insight into the generation of antibodies in severe infection [22]. Given the ease and speed of performing our serology assays, these could be included in a clinical stratification for intervention in inpatient COVID-19 infections.

The higher levels of production of anti-viral antibodies during SARS-CoV-2 infections that subsequently resolved may be indication of an immune response that is skewed away from a pathogenic inflammatory response. Pro-inflammatory cytokines and T cell responses have been associated with poor clinical outcome in COVID-19 [25, 26], but IL4 and IL5 have also been found to be elevated in severe infection [27]. A unique, complex dysregulation of the immune response has been described in severe COVID-19 disease [28]. The relationship between the early events in severe disease clearly requires further investigation.

A further major finding was the discovery of differing patterns of anti-S1 subunit antibodies and the neutralizing potency of sera between different patients. Neutralisation (and enhancement) of virus entry was observed in some serum samples where there was no detectable anti-S1 (Figure 4C). There must therefore be specific components of the polyclonal anti-spike antibody response generated by SARS-CoV-2 infection that are not detected in S1-based antibody assays. The sera used were heat-inactivated to prevent complement-mediated inhibition and did not neutralize non-SARS-CoV-2 pseudotypes. Lower dilutions of inactivated sera were cytotoxic, due to the presence of Triton X-100, but our optimised dilution of 1:300 (0.0033% Triton X-100) was tolerated by the target cells, providing resolvable inhibition of SARS-CoV-2 entry. As such, we conclude that the inhibition was specific to the antibodies present in each sample.

Enhancement of virus entry was observed for some serum samples, mostly in samples that had very low reactivity in the anti-S1 assay (Figure 4C). This finding is particularly important for designing a clinically useful vaccine; if virus enhancing antibodies are generated by immunization, it is likely that protection will not be provided. Current early data from ongoing vaccine trials report high titres of neutralizing antibodies [29, 30]. Antibody-dependent enhancement has been described for viruses from different families, including coronaviruses such as feline infectious peritonitis virus (FIPV) [31] and SARS-CoV-1 [32]. ADE has been proposed to make a possible contribution to pathogenesis of COVID-19 [33]. Our data support the need to investigate this phenomenon in more detail, interrogating the dose-dependent nature of neutralization and enhancement [32].

For the first time, we also interrogated the antibody response in patients known to be infected with different SARS-CoV-2 variants. The spike 614G polymorphism is unusual as it has progressively spread throughout Europe, becoming the major variant [13]. In our cohort, the 614G variant was present in 55% of the samples analysed. While the antigens used in our ELISA and neutralization assays possessed the 614D variant, reactivity and neutralization was greater for sera isolated from infections with the 614G viruses. This implicates this variant in increased immunogenicity. Given the increased stability of this variant [34], antibodies elicited by viruses with 614G may have greater binding ability for correctly folded spike protein, resulting in greater neutralizing potency. Antibodies directed to the 614D variant, which if more likely to be shed from the surface of a virus particle, may recognise epitopes that do not result in effective neutralization, and may be more likely to enhance infection. While this cannot directly account for the greater reactivity to the nucleocapsid protein observed in these patients, the viruses possessing 614G might be globally more antigenic. It is also plausible that greater anti-nucleocapsid reactivity in infections possessing the 614G variant is an indirect indication of greater viral load at the site of infection [13]. This finding is also important for vaccine development; the increased neutralization by antibodies from infections with 614G may indicate that this variant will make a more potent immunogen. It will be important to further profile the specificity of individual antibodies in different infections to identify the determinants of neutralization of SARS-CoV-2.

Whilst some studies on hospitalised individuals have failed to show an association between antibody titre and clinical severity of disease [35], analysis of the assays used in those reports suggest that saturating amounts of antibodies (that is, low serum dilutions) were used, which were unable to resolve the differences in overall titres reported here. Also, our study focussed on mainly Caucasian, elderly inpatients. Differences in ethnic background, and other variables, may have a role in determining the quality of the antibody responses in SARS-CoV-2 infected patients.

To our knowledge this study provides the first evidence that a potent antibody response early in infection may contribute to clearance of infection and improved prognosis, even when disease symptoms are severe. While there are limitations on interpretation of our data given the retrospective sampling approach we have adopted, further analysis of the potency of neutralizing antibody responses, and the potential for infection enhancement are warranted. The data presented here also supports the rationale for use of optimised vaccines that generate antibodies directed to the spike and nucleocapsid genes and elicit potently neutralizing antibodies. The use of convalescent sera as a therapeutic approach [36] may also provide sufficient potency to contribute to resolution of infection, if potently neutralizing sera are specifically selected.

## Data Availability

All raw data is available on request

## Acknowledgements

This project received funding from the University of Nottingham Campaign and Alumni Relations Office and an MRC Confidence in Concept award. Whole genome sequencing of SARS-CoV-2 was funded by COG-UK; COG-UK is supported by funding from the Medical Research Council (MRC) part of UK Research & Innovation (UKRI), the National Institute of Health Research (NIHR) and Genome Research Limited, operating as the Wellcome Sanger Institute.

## Author contributions

Conceived and designed the study: P.J.T., J.K.B., W.L.I., A.W.T.; Performed experiments: P.J.T., R.A.U, L.F., C.P.M. J.G.C., N.H., M.C, C.M., A.W.T.; Analysed data P.J.T., R.A.U., N.G., T.T., M.L., F.S., A.W.T. Provided clinical data: B.J.T., D.H., G.C., T.B., W.L.I,; Provided essential reagents: N.T.; Wrote manuscript: A.W.T.; Edited manuscript: P.J.T, J.K.B., W.L.I, A.W.T.; Reviewed and agreed final version: all authors; Secured funding; P.J.T., J.K.B., A.W.T.

